# Assessing the impact of interventions on the major Omicron BA.2 outbreak in spring 2022 in Shanghai

**DOI:** 10.1101/2024.01.08.24300974

**Authors:** Hengcong Liu, Jun Cai, Jiaxin Zhou, Xiangyanyu Xu, Marco Ajelli, Hongjie Yu

**Affiliations:** School of Public Health, Fudan University, Key Laboratory of Public Health Safety, Ministry of Education, Shanghai, China; Laboratory for Computational Epidemiology and Public Health, Department of Epidemiology and Biostatistics, Indiana University School of Public Health, Bloomington, IN, USA; Shanghai Institute of Infectious Disease and Biosecurity, School of Public Health, Fudan University, Shanghai, China

## Abstract

**Background:** Shanghai experienced a significant surge in Omicron BA.2 infections from March to June 2022. In addition to the standard interventions in place at that time, additional interventions were implemented in response to the outbreak. However, the impact of these interventions on BA.2 transmission remains unclear.

**Methods:** We systematically collected data on the daily number of newly reported infections during this wave and utilized a Bayesian approach to estimate the daily effective reproduction number. Data on public health responses were retrieved from the Oxford COVID-19 Government Response Tracker and served as a proxy for the interventions implemented during this outbreak. Using a log-linear regression model, we assessed the impact of these interventions on the reproduction number. Furthermore, we developed a mathematical model of BA.2 transmission. By combining the estimated effect of the interventions from the regression model and the transmission model, we estimated the number of infections and deaths averted by the implemented interventions.

**Results:** We found a negative association (-0.0069, 95% CI: -0.0096 to -0.0045) between the level of interventions and the number of infections. If interventions did not ramp up during the outbreak, we estimated that the number of infections and deaths would have increased by 22.6% (95% CI: 22.4-22.8%), leading to a total of 768,576 (95% CI: 768,021-769,107) infections and 722 (95% CI: 722-723) deaths. If no interventions were deployed during the outbreak, we estimated that the number of infections and deaths would have increased by 46.0% (95% CI: 45.8-46.2%), leading to a total of 915,099 (95% CI: 914,639-915,518) infections and 860 (95% CI: 860-861) deaths.

**Conclusion:** Our findings suggest that the interventions adopted during the Omicron BA.2 outbreak in spring 2022 in Shanghai were effective in reducing SARS-CoV-2 transmission and disease burden. Our findings emphasize the importance of non-pharmacological interventions in controlling quick surges of cases during epidemic outbreaks.

## Background

China experienced multiple localized Omicron outbreaks in the first half of 2022 [1]. Specifically, Shanghai underwent a major outbreak of the Omicron BA.2 variant in March 2022, resulting in a total of 0.62 million reported infections (∼2.4% of the Shanghai population) and 588 deaths [2]. In response to this outbreak, a set of additional interventions were implemented, including district-wide and city-wide mass nucleic acid screening, as well as lockdown measures [3]. Moreover, Hong Kong faced its fifth wave of coronavirus disease 2019 (COVID-19) in late December 2021, leading to over 3.0 million infections [4], affecting a substantial 41.1% of the Hong Kong population. To safeguard people’s health and protect the healthcare system from collapse, mitigation strategies such as rapid antigen tests (RATs) [5] and the “StayHomeSafe” scheme [6] were deployed. These measures have been demonstrated to significantly contribute to reducing the reproduction number and easing the isolation and treatment burden of hospitals [7]. However, the quantitative impact of these interventions on BA.2 transmission in Shanghai has yet to be quantified.

In this study, we utilized a log-linear regression model to evaluate the impact of these interventions on the reproduction number. Furthermore, we employed a stochastic susceptible-latent-infectious-recovered (SLIR) model to simulate the transmission of BA.2 in Shanghai. The objective of this study is to assess the effect of these interventions on the disease burden and estimate the number of averted infections and deaths under counterfactual scenarios on alternative interventions.

## Methods

### Data sources

For the entire duration of the Omicron BA.2 wave in Spring 2022 in Shanghai, we systematically collected data on the daily number of newly reported COVID-19 infections from March 1 to June 1, 2022 (Supplementary Table 1) [2].

### Public health response

We downloaded public health response data from the Oxford COVID-19 Government Response Tracker [8]. Changes in policy indicators related to containment and closure policies (C1-C8) and health system policies (H1-H8) in Shanghai from February to July 2022 are illustrated in Supplementary Figure 1. The containment and health index (denoted as *C_t_*) served as an index grouping these two families of policy indicators. Before the start of the Omicron BA.2 wave, Shanghai maintained a baseline level of non-pharmacological interventions to prevent potential outbreak of COVID-19. Between March and June 2022, additional interventions, including district-wide and city-wide mass nucleic acid screening and lockdown, were implemented in response to the outbreak. The majority of the containment measures (such as lockdowns) were lifted at the end of the outbreak (Figure 1 and Supplementary Table 2).

**Figure 1.**
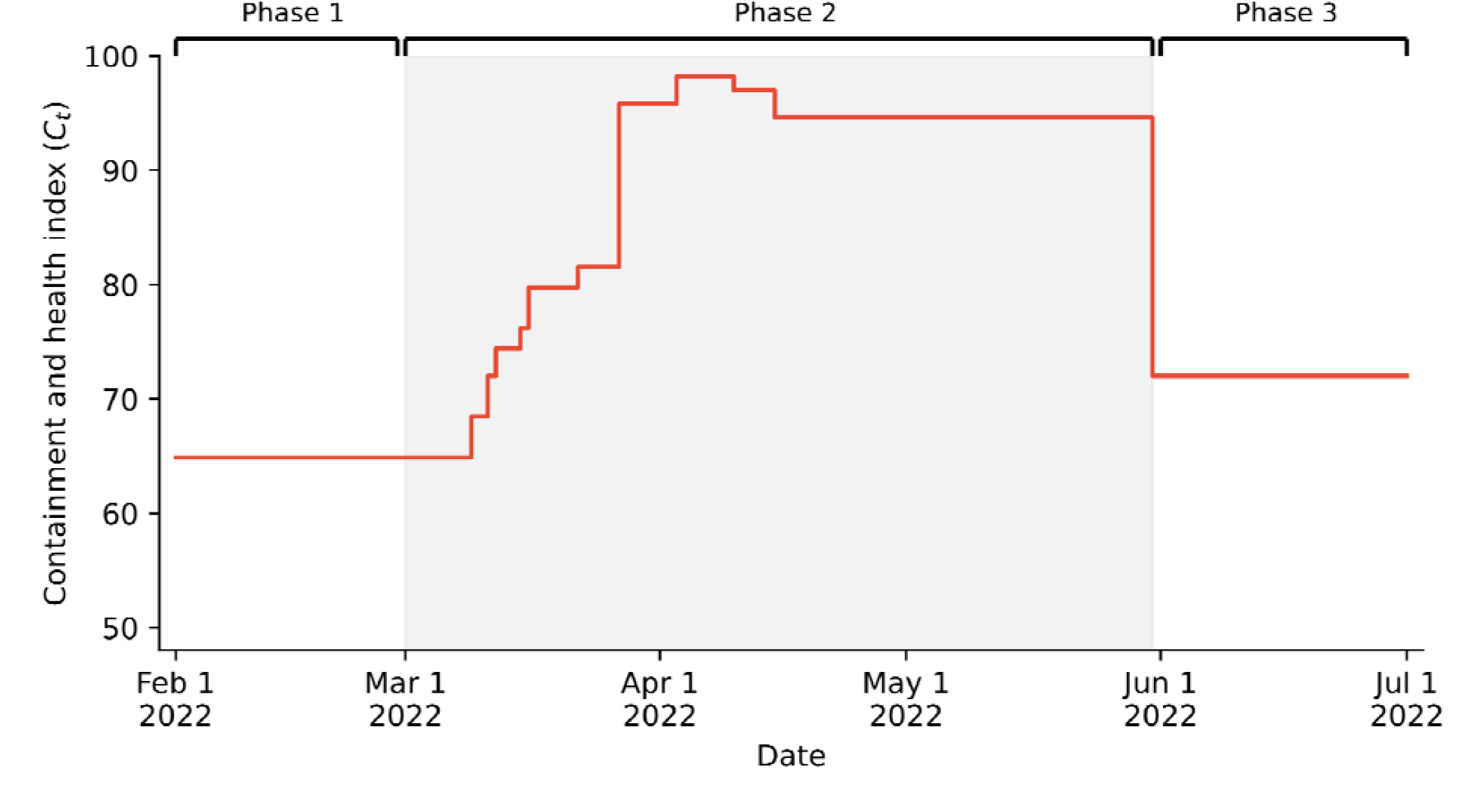
Containment and health index () over time during the Omicron BA.2 outbreak in Shanghai. Phase 1 (Pre-March 2022): Implementation of baseline interventions to prevent potential COVID-19 outbreaks. Phase 2 (March to June 2022, shaded grey area): Implementation of baseline and additional interventions in response to the Omicron BA.2 outbreak. Phase 3 (Post-June 2022): Easing of containment and closure policies.

### Effective reproduction number

We applied a Bayesian approach to estimate the real-time daily effective reproduction number (denoted as 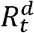) [9]. This method relies on the time series of the daily number of new infections and knowledge of the distribution of the generation time. Since our dataset provided only the official reporting date, we generated the date of sample collection for each infection using a Poisson distribution with the mean equal to the delay between the date of sample collection and the reporting date. In our main analysis, we considered delays of 2 days, 3 days, and 2 days between the date of sample collection and reporting for periods before March 15, between March 16 and May 14, and after May 15, respectively, based previous findings [3]. Additionally, a 4-day delay for the period between March 16 and May 14 was considered as a sensitivity analysis. This estimated sampling date was assumed to approximate the date of symptom onset. The generation time was assumed to follow a gamma distribution of mean 2.7 days (shape: 3.25, scale: 0.84), in agreement with the mean serial interval estimated in reference [10]. As a sensitivity analysis, we considered a gamma distribution of mean 3.3 days (shape: 1.89, scale: 1.75), in agreement with the mean serial interval estimated in reference [11]. The weekly effective reproduction number (denoted as 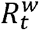) was derived as the weekly geometric mean of 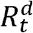.

### Log-linear regression model

Following the approach presented in reference [12]. we used a regression model to establish the relationship among 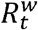, the depletion of susceptible individuals, and implemented interventions *C_t_*. The log-linear regression model is expressed as:

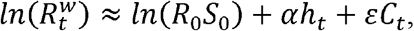

where:

- *R*_0_ represents the basic reproduction number,
- *S*_0_ represents the initial fraction of susceptible individuals in the population,
- *h_t_* represents the observed cumulative incidence of infections up to week *t*-1,
- *α* represents the rate at which susceptible individuals are depleted,
- *ε* represents the coefficient measuring the effect of the implemented interventions.

R-squared was calculated to measure the goodness of fit of the regression model. In the main analysis, we analyzed an 11-week dataset of 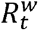, centered on the peak incidence of reported COVID-19 infections (April 9, 2022). This dataset covered the period from March 2 to May 17, 2022. Additionally, we performed a sensitivity analysis by adjusting the duration of the dataset to 10 or 12 weeks.

### SARS-CoV-2 transmission model

We utilized the standard SLIR model to simulate the transmission of BA.2 in Shanghai (Supplementary Method). This model classified the population into four epidemiological categories: susceptible, latent, infectious, and recovered. Upon infection, susceptible individuals enter a latent compartment and become infectious after a latent period of 1.0 days[13]. Infectious individuals could transmit the virus to susceptible individuals during an infectious period of 1.7 days, after which they naturally recover. Since latent and infectious periods are both exponentially distributed, their sum corresponds to the generation time of average 2.7 days [14]. We modeled the transitions between compartments using a stochastic chain binomial process [15, 16]. Given the limited protection against infection provided by the inactivated vaccine and the rapid decline in immunity [17], we assumed that the population was fully susceptible to infection during the analyzed outbreak.

Using the estimated coefficients in the regression model, we determined the initial net reproduction number (denoted as *R_e_*) as 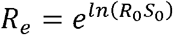. The transmission rate in absence of interventions (denoted as *β*_0_) was estimated following its relationship with 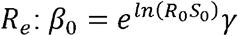, where *γ* represents the recovery rate. The number of reported infections was assumed to be proportional to the number of infected individuals, and the seeding date was set at March 1, 2022. To calibrate the model, we fitted it to the time series of cumulative infections, identifying the number of initial infectious seeds that best matched the observed data.

The baseline scenario (denoted as S0) corresponds to the real-world situation, and simulations were carried out using the observed time series of interventions (*C_t_*) and the estimated real-time transmission rate 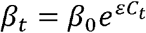. Then, we reconstructed the transmission rate *β_t_* by varying *C_t_* values to simulate the transmission of BA.2 under two counterfactual scenarios. Counterfactual scenario 1 (denoted as S1) assumes that the level of interventions remained constant at the level observed before the start of the outbreak, namely 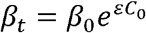 . In counterfactual scenario 2 (denoted as S2), no interventions were implemented, namely the values of *C_t_* were set to zero throughout the entire outbreak and *β_t_ = β_0_*. Given the number of infections projected by the transmission model, the overall crude infection-fatality ratio (0.094% [2]) was used to estimate the number of deaths.

## Results

A total of 626,806 infections were officially reported in Shanghai between March and June 2022 (Figure 2A and Supplementary Figure 2). In response to this outbreak, additional interventions were implemented. 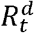 initially fluctuated around 2 and gradually decreased due to district-wide mass nucleic acid screening and lockdown. Subsequently, there was a rapid drop below the epidemic threshold following the implementation of city-wide mass nucleic acid screening and lockdown (Figure 2B). Consistent findings were observed when considering a 4-day delay between the sampling date and reporting date in the period from March 16 to May 14, 2022, and using a gamma-distributed generation time with a mean of 3.3 days (Supplementary Figure 3).

**Figure 2.**
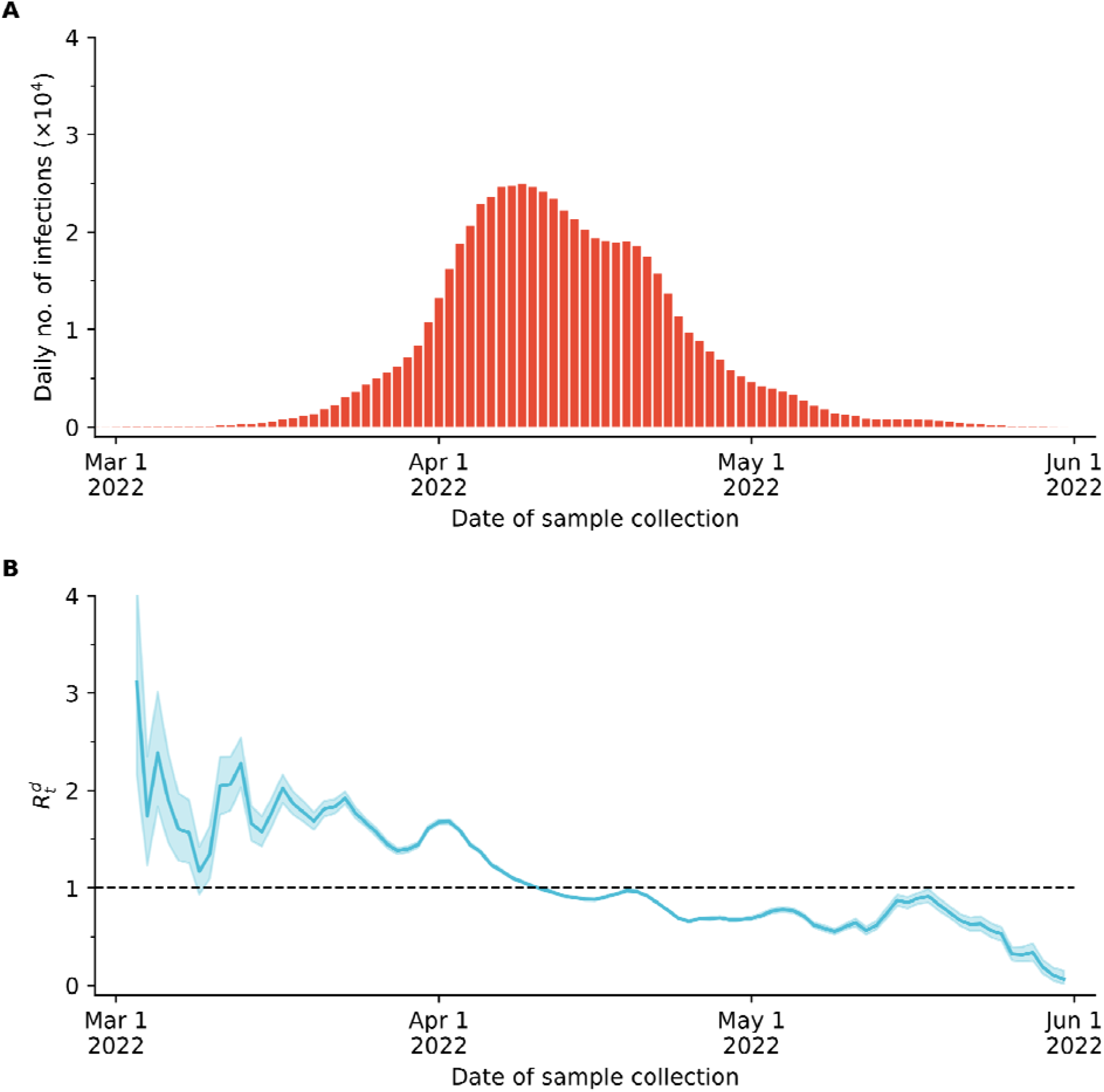
Epidemic curve and estimated during the Omicron BA.2 outbreak in Shanghai. **A** Daily number of newly reported infections by date of sample collection. **B** Daily effective reproduction number as estimated from our Bayesian approach. Lines and shaded areas: mean and 95% CI of 100 simulations. The horizontal line represents the epidemic threshold.

We applied a log-linear regression model to assess the impact of interventions on the reproduction number (Figure 3A). The parameter *ε* was estimated to be -0.0069 (95% CI: - 0.0096, -0.0045). This negative value indicates that the implementation of interventions effectively reduced the reproduction number. The estimated parameters of the regression model are reported in Supplementary Table 3. Shortening or lengthening the duration of dataset to 10 or 12 weeks had little effect on model outcomes (Supplementary Figure 4-5).

**Figure 3.**
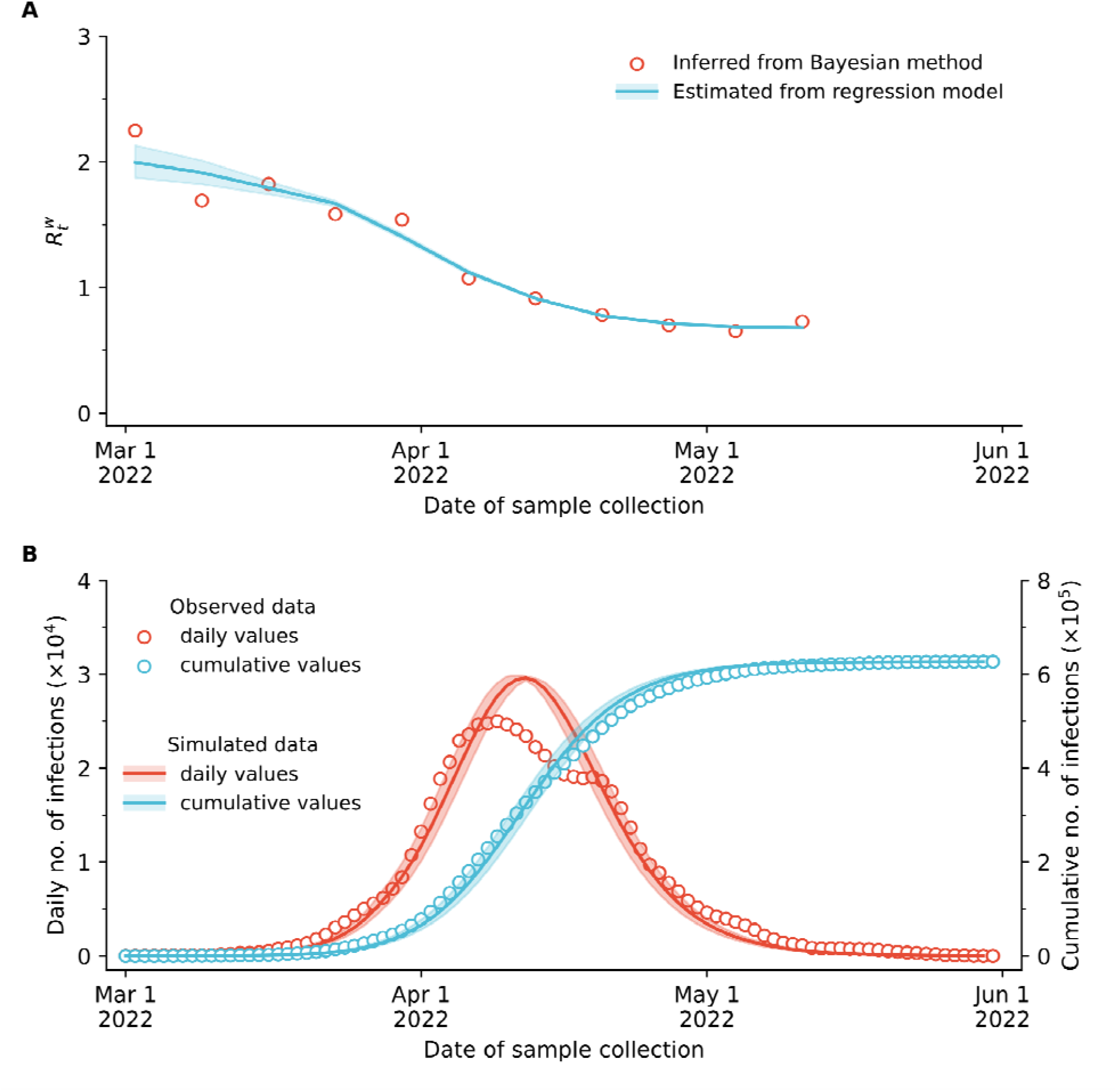
Fit of the weekly effective reproduction number and epidemic curves. **A** Weekly effective reproduction number as estimated from our Bayesian approach (red circles) and as estimated from our regression model (blue line). Blue line and shaded areas: mean and 95% CI of 100 simulations. **B** Daily number of new and cumulative infections as observed in the data (red or blue circles) and as simulated by the SLIR model (red or blue lines). Lines and shaded areas: mean and 95% CI of 100 simulations.

We then fit the transmission model to the time series of cumulative infections. The best fit was obtained by setting initial infectious seeds to 230, which projected 626,858 (95% CI: 625,959-627,674) infections and 589 (95% CI: 588-590) deaths (Figure 3B). Using the calibrated model, we assessed the impact of interventions on the disease burden (Figure 4). In counterfactual scenario 1, the estimated cumulative number of infections and deaths were 768,576 (95% CI: 768,021-769,107) infections and 722 (95% CI: 722-723), respectively. Compared to the baseline scenario, the absence of additional interventions would have resulted in a 22.6% (95% CI: 22.4-22.8%) increase in disease both in the number of infections and deaths. Counterfactual scenario 2 projected a total of 915,099 (95% CI: 914,639-915,518) infections and 860 (95% CI: 860-861) deaths. This suggests that if no interventions were deployed the disease burden would have increased by 46.0% (95% CI: 45.8-46.2%) as compared to the baseline scenario.

**Figure 4.**
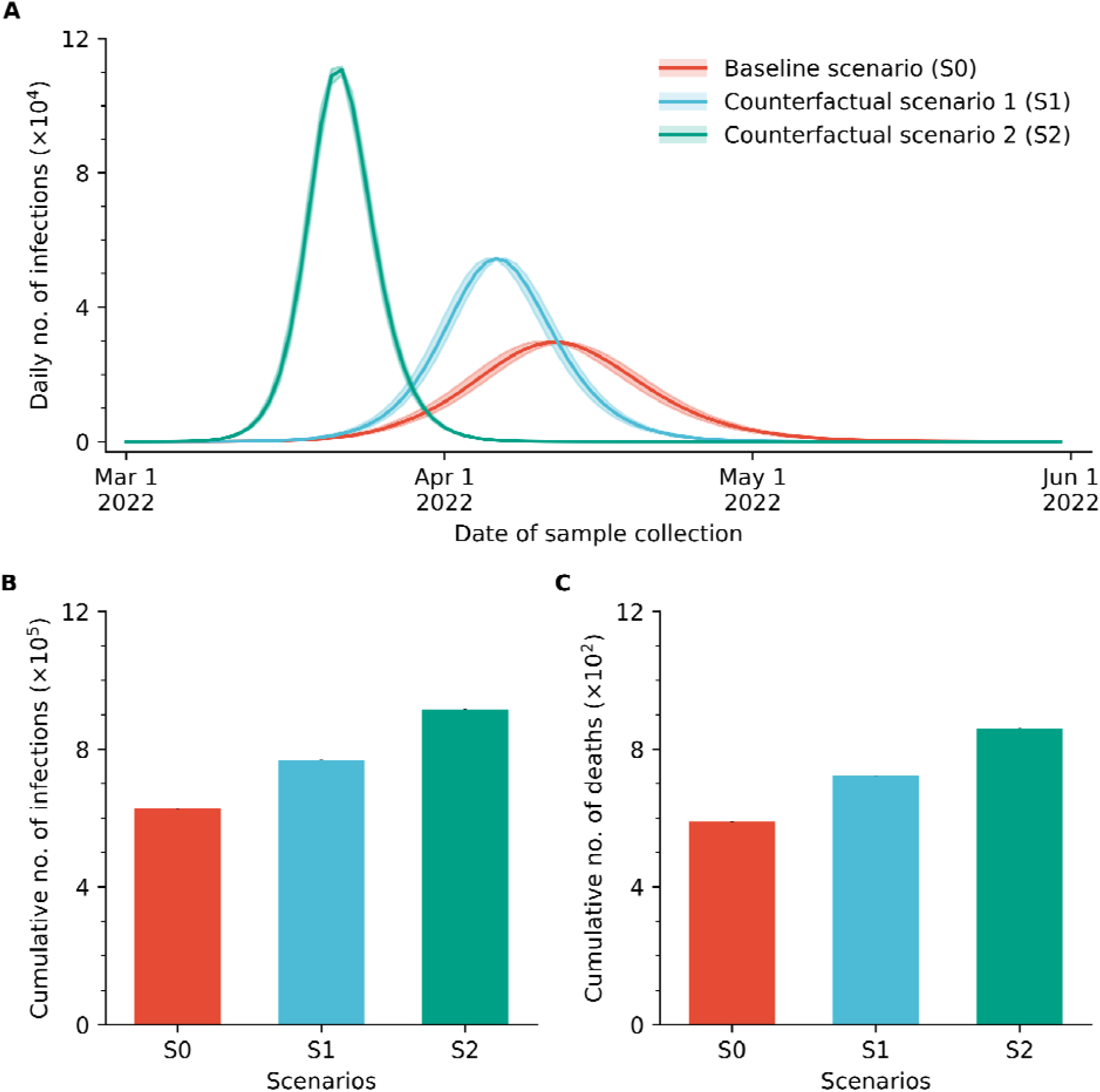
Projected disease burden under baseline and counterfactual scenarios. **A** Daily number of new infections over time under different scenarios. Baseline scenario (S0): simulations are carried out using the observed time series of interventions. Counterfactual scenario 1 (S1): simulations are carried out maintaining constant pre-outbreak interventions. Counterfactual scenario 2 (S2): no interventions. Lines and shaded areas: mean and 95% CI of 100 simulations. **B** Cumulative number of infections under the three scenarios used in panel A. **C** Cumulative number of deaths under the three scenarios used in panel A.

## Discussion

In this study, we used a combination of log-linear regression and mechanistic modeling to evaluate the impact of interventions on the Omicron BA.2 outbreak in spring 2022 in Shanghai, China. We estimated that escalation of non-pharmacological interventions in response to the outbreak decreased the disease burden by 22.6% (95% CI: 22.4-22.8%). Moreover, we estimated that in the absence of any intervention, the disease burden would have increased by 46.0% (95% CI: 45.8-46.2%).

Throughout the Shanghai Omicron BA.2 wave, most of the infectious individuals were identifies though multiple rounds of mass nucleic acid screening characterized by high test sensitivity (pool size of 5-20 samples were commonly tested in the outbreak, which would lead to a detection sensitivity of 84.7-95.3% [18]). Subsequently, these individuals were either isolated in the designated hospitals or dedicated isolation facilities, or they were quarantined at home. This approach contributed for the interruption the transmission process of infectious individuals, contributing to explain the high effectiveness of the interventions estimated in this study.

Our study has several limitations that should be considered. First, the estimation of the effective reproduction number over time is based on the date of sample collections inferred from the date of reporting. Although this is not ideal, previous studies have used a similar approach [3, 19]. Moreover, it is important to stress that multiple rounds of mass screenings were implemented during the analyzed outbreak, greatly reducing the delay between infection and reporting dates. Finally, we also conducted a sensitivity analysis with different reporting delays and found consistent results. Second, the adopted interventions have likely contributed to the shortening of the duration generation time as compared to the intrinsic generation time of Omicron [20]. Since no estimates of the realized generation time during the Omicron BA.2 outbreak in Shanghai is available in the literature, we relied on estimates of the serial interval available for an outbreak in Hong Kong that was treated with strict interventions [10]. To assess the robustness of our findings, we conducted a sensitivity analysis assuming a longer generation time and found similar results. Third, we estimated the overall effect of the interventions adopted during the Shanghai outbreak. This analysis was based on the containment and health index from the Oxford COVID-19 Government Response Tracker was used as a proxy of the intensity of the interventions implemented during the outbreak, which has been widely adopted in previous studies [21-23]. Further analyses are warranted to investigate the impact of single interventions separately.

In conclusion, our study estimated the effect of the interventions adopted during the Omicron BA.2 outbreak in spring 2022 in Shanghai in reducing SARS-C-V-2 transmission, measured as effective reproduction number, and disease burdens, measured as number of infections and deaths. Our findings emphasize the importance of non-pharmacological interventions in controlling quick surges of cases during epidemic outbreaks.

## Data Availability

The code and data used to conduct these analyses are available through a request.

## Declarations

### Authors’ contributions

H.L., M.A. and H.Y. conceived and designed the study. H.Y. and M.A. supervised the study. H.L. designed and developed the model and analyzed the model outputs. H.L., J.Z., J.C. and M. A. prepared the first draft of the manuscript. H.L. and X.X. prepared the tables and figures. All authors contributed to review and revision, approved the final manuscript as submitted and agree to be accountable for all aspects of the work.

### Competing interests

H.Y. has received research funding from Sanofi Pasteur, GlaxoSmithKline, Yichang HEC Changjiang Pharmaceutical Company, and Shanghai Roche Pharmaceutical Company, and SINOVAC Biotech Ltd. None of these research funding is related to this work. All other authors report no competing interests.

### Funding

The study was supported by Shanghai Municipal Science and Technology Major Project (No. ZD2021CY001 to H.Y.) and the Young Scientists Fund of the National Natural Science Foundation of China (No. 82304199 to J.C.). The funders had no role in study design, data collection, data analysis, data interpretation or writing of the report.

### Availability of data and materials

The code and data used to conduct these analyses are available through a request.

### Ethics approval and consent to participate

Not applicable.

## Acknowledgements

Not applicable.

## Consent for publication

Not applicable.

## Notes

### Competing Interest Statement

The authors have declared no competing interest.

### Author Declarations

The study used ONLY openly available human data that were originally located at official reports.

## References

1. National Health Commission of the People’s Republic of China. Update on COVID-19. [cited 2023 October 30]; Available from: http://www.nhc.gov.cn/xcs/yqtb/list_gzbd.shtml.

2. The State Council Information Office of the People’s Republic China. The press conference of COVID-19 in Shanghai. [cited 2023 October 30]; Available from: http://www.scio.gov.cn/xwfb/dfxwfb/gssfbh/sh_13834/.

3. Chen, Z., et al., Epidemiological characteristics and transmission dynamics of the outbreak caused by the SARS-CoV-2 Omicron variant in Shanghai, China: A descriptive study. Lancet Reg Health West Pac, 2022. 29: p. 100592.

4. Centre for Health Protection of the Department of Health; and the Hospital Authority. Statistics on the fifth wave of COVID-19. [cited 2023 October 30]; Available from: https://www.coronavirus.gov.hk/pdf/5th_wave_statistics/5th_wave_statistics_20230727.pdf.

5. The Government of the Hong Kong Special Administrative Region. Government continues adopting risk-based testing strategy. [cited 2023 October 30]; Available from: https://www.info.gov.hk/gia/general/202202/25/P2022022500816.htm.

6. The Government of the Hong Kong Special Administrative Region. “StayHomeSafe” Scheme launched. [cited 2023 October 30]; Available from: https://www.info.gov.hk/gia/general/202202/08/P2022020800726.htm.

7. Du, Z., L. Tian, and D.-Y. Jin, Understanding the impact of rapid antigen tests on SARS-CoV-2 transmission in the fifth wave of COVID-19 in Hong Kong in early 2022. Emerging Microbes & Infections, 2022. 11(1): p. 1394–1401.

8. Hale, T., et al., A global panel database of pandemic policies (Oxford COVID-19 Government Response Tracker). Nature Human Behaviour, 2021. 5(4): p. 529–538.

9. Liu, Q.H., et al., Measurability of the epidemic reproduction number in data-driven contact networks. Proc Natl Acad Sci U S A, 2018. 115(50): p. 12680–12685.

10. Mefsin, Y.M., et al., Epidemiology of Infections with SARS-CoV-2 Omicron BA.2 Variant, Hong Kong, January–March 2022. Emerging Infectious Diseases, 2022. 28(9): p. 1856–1858.

11. Backer, J.A., et al., Shorter serial intervals in SARS-CoV-2 cases with Omicron BA.1 variant compared with Delta variant, the Netherlands, 13 to 26 December 2021. Eurosurveillance, 2022. 27(6): p. 2200042.

12. te Beest, D.E., et al., Driving Factors of Influenza Transmission in the Netherlands. American Journal of Epidemiology, 2013. 178(9): p. 1469–1477.

13. Leung, K., et al., Estimating the transmission dynamics of SARS-CoV-2 Omicron BF.7 in Beijing after the adjustment of zero-COVID policy in November - December 2022. Nat Med, 2023.

14. Wallinga, J. and M. Lipsitch, How generation intervals shape the relationship between growth rates and reproductive numbers. Proc Biol Sci, 2007. 274(1609): p. 599–604.

15. Cai, J., et al., Modeling transmission of SARS-CoV-2 Omicron in China. Nat Med, 2022. 28(7): p. 1468–1475.

16. Liu, H., et al., Projecting the potential impact of an Omicron XBB.1.5 wave in Shanghai, China. medRxiv, 2023: p. 2023.05.10.23289761.

17. Huang, Z., et al., Effectiveness of inactivated and Ad5-nCoV COVID-19 vaccines against SARS-CoV-2 Omicron BA. 2 variant infection, severe illness, and death. BMC Medicine, 2022. 20(1).

18. Yelin, I., et al., Evaluation of COVID-19 RT-qPCR Test in Multi sample Pools. Clin Infect Dis, 2020. 71(16): p. 2073–2078.

19. Abbott, S., et al., Estimating the time-varying reproduction number of SARS-CoV-2 using national and subnational case counts [version 2; peer review: 1 approved, 1 approved with reservations]. Wellcome Open Research, 2020. 5(112).

20. Manica, M., et al., Intrinsic generation time of the SARS-CoV-2 Omicron variant: An observational study of household transmission. Lancet Reg Health Eur, 2022. 19: p. 100446.

21. Ge, Y., et al., Untangling the changing impact of non-pharmaceutical interventions and vaccination on European COVID-19 trajectories. Nature Communications, 2022. 13(1): p. 3106.

22. Zhang, X., et al., Assessing the impact of COVID-19 interventions on influenza-like illness in Beijing and Hong Kong: an observational and modeling study. Infectious Diseases of Poverty, 2023. 12(1): p. 11.

23. García-García, D., et al., Assessing the effect of non-pharmaceutical interventions on COVID-19 transmission in Spain, 30 August 2020 to 31 January 2021. Eurosurveillance, 2022. 27(19): p. 2100869.

