## Supplemental Table 1-3, Figure 1-5 for "Assessing the impact of interventions on the major Omicron BA.2 outbreak in spring 2022 in Shanghai"

### Supplementary method

We employed a stochastic susceptible-latent-infectious-recovered (SLIR) model to simulate the transmission of Omicron BA.2 in Shanghai. The transmission model can be described by the following system of differential equations:

$$\frac{dS(t)}{dt}=-\lambda_{t}S(t)$$

$$\frac{dL(t)}{dt}=\lambda_{t}S(t)-\sigma L\left( t \right)$$

$$\frac{dI(t)}{dt}=\sigma L\left( t \right)-\gamma I\left( t \right)$$

$$\frac{dR(t)}{dt}=\gamma I\left( t \right)$$

where:

- $S(t)$ represents the number of susceptible individuals at time $t$.
- $L(t)$ represents the number of latent individuals at time $t$.
- $I(t)$ represents the number of infectious individuals at time $t$.
- $R(t)$ represents the number of recovered individuals at time $t$.
- $\lambda_{t}$ represents the transmission rate.
- $1/\sigma$ represents the latent period.
- $1/\gamma$ represents the infectious period.

Transitions between compartments are modelled through a stochastic chain binomial process. Specifically, the number of individuals transitioning from the susceptible to the latent compartment is determined by a binomial distribution $Bin(S(t), \lambda(t))$.

The time-dependent force of infection $\lambda_{t}$ is defined as:

$$\lambda_{t}=\beta_{0}e^{\varepsilon C_{t}}\frac{I(t)}{N}$$

where:

- $\beta_{0}$ is the transmission risk in the absence of interventions.
- $\varepsilon$ is the coefficient that measures the effect of interventions in the regression model.
- $C_{t}$ is the proxy that represents the interventions implemented during the Shanghai Omicron BA.2 wave.
- $N$ is total number of Shanghai population.

### Supplementary results

#### Supplementary Table 1. Epidemic data during the Shanghai Omicron BA.2 wave

| Date | Symptomatic cases | Asymptomatic cases | Infections |
| --- | --- | --- | --- |
| 2022/3/1 | 1 | 1 | 2 |
| 2022/3/2 | 3 | 5 | 8 |
| 2022/3/3 | 2 | 14 | 16 |
| 2022/3/4 | 3 | 16 | 19 |
| 2022/3/5 | 0 | 28 | 28 |
| 2022/3/6 | 3 | 45 | 48 |
| 2022/3/7 | 4 | 51 | 55 |
| 2022/3/8 | 3 | 62 | 65 |
| 2022/3/9 | 4 | 76 | 80 |
| 2022/3/10 | 11 | 64 | 75 |
| 2022/3/11 | 5 | 78 | 83 |
| 2022/3/12 | 1 | 64 | 65 |
| 2022/3/13 | 39 | 128 | 167 |
| 2022/3/14 | 9 | 130 | 139 |
| 2022/3/15 | 5 | 197 | 202 |
| 2022/3/16 | 7 | 150 | 157 |
| 2022/3/17 | 57 | 203 | 260 |
| 2022/3/18 | 8 | 366 | 374 |
| 2022/3/19 | 11 | 492 | 503 |
| 2022/3/20 | 24 | 734 | 758 |
| 2022/3/21 | 31 | 865 | 896 |
| 2022/3/22 | 4 | 977 | 981 |
| 2022/3/23 | 4 | 979 | 983 |
| 2022/3/24 | 29 | 1,580 | 1,609 |
| 2022/3/25 | 33 | 2,231 | 2,264 |
| 2022/3/26 | 45 | 2,631 | 2,676 |
| 2022/3/27 | 50 | 3,450 | 3,500 |
| 2022/3/28 | 75 | 4,381 | 4,456 |
| 2022/3/29 | 308 | 5,656 | 5,964 |
| 2022/3/30 | 339 | 5,298 | 5,637 |
| 2022/3/31 | 338 | 4,144 | 4,482 |
| 2022/4/1 | 258 | 6,051 | 6,309 |
| 2022/4/2 | 365 | 7,788 | 8,153 |
| 2022/4/3 | 354 | 8,581 | 8,935 |
| 2022/4/4 | 264 | 13,086 | 13,350 |
| 2022/4/5 | 271 | 16,766 | 17,037 |
| 2022/4/6 | 307 | 19,660 | 19,967 |
| 2022/4/7 | 501 | 20,398 | 20,899 |
| 2022/4/8 | 595 | 22,609 | 23,204 |
| 2022/4/9 | 815 | 23,937 | 24,752 |
| 2022/4/10 | 867 | 25,173 | 26,040 |
| 2022/4/11 | 721 | 22,348 | 23,069 |
| 2022/4/12 | 1,166 | 25,141 | 26,307 |
| 2022/4/13 | 2,459 | 25,146 | 27,605 |
| 2022/4/14 | 2,893 | 19,872 | 22,765 |
| 2022/4/15 | 2,668 | 19,923 | 22,591 |
| 2022/4/16 | 2,061 | 21,582 | 23,643 |
| 2022/4/17 | 1,564 | 19,831 | 21,395 |
| 2022/4/18 | 2,110 | 17,332 | 19,442 |
| 2022/4/19 | 1,961 | 16,407 | 18,368 |
| 2022/4/20 | 2,175 | 15,861 | 18,036 |
| 2022/4/21 | 1,788 | 15,698 | 17,486 |
| 2022/4/22 | 1,616 | 20,634 | 22,250 |
| 2022/4/23 | 860 | 19,657 | 20,517 |
| 2022/4/24 | 1,626 | 16,983 | 18,609 |
| 2022/4/25 | 693 | 15,319 | 16,012 |
| 2022/4/26 | 353 | 11,956 | 12,309 |
| 2022/4/27 | 434 | 9,330 | 9,764 |
| 2022/4/28 | 425 | 9,545 | 9,970 |
| 2022/4/29 | 264 | 8,932 | 9,196 |
| 2022/4/30 | 105 | 7,084 | 7,189 |
| 2022/5/1 | 198 | 6,606 | 6,804 |
| 2022/5/2 | 119 | 5,395 | 5,514 |
| 2022/5/3 | 109 | 4,722 | 4,831 |
| 2022/5/4 | 76 | 4,390 | 4,466 |
| 2022/5/5 | 64 | 4,024 | 4,088 |
| 2022/5/6 | 78 | 3,961 | 4,039 |
| 2022/5/7 | 80 | 3,760 | 3,840 |
| 2022/5/8 | 92 | 3,625 | 3,717 |
| 2022/5/9 | 78 | 2,780 | 2,858 |
| 2022/5/10 | 30 | 1,259 | 1,289 |
| 2022/5/11 | 38 | 1,305 | 1,343 |
| 2022/5/12 | 60 | 1,869 | 1,929 |
| 2022/5/13 | 54 | 1,487 | 1,541 |
| 2022/5/14 | 55 | 1,203 | 1,258 |
| 2022/5/15 | 27 | 869 | 896 |
| 2022/5/16 | 31 | 746 | 777 |
| 2022/5/17 | 40 | 759 | 799 |
| 2022/5/18 | 34 | 637 | 671 |
| 2022/5/19 | 17 | 770 | 787 |
| 2022/5/20 | 35 | 784 | 819 |
| 2022/5/21 | 23 | 570 | 593 |
| 2022/5/22 | 25 | 503 | 528 |
| 2022/5/23 | 19 | 422 | 441 |
| 2022/5/24 | 12 | 343 | 355 |
| 2022/5/25 | 17 | 290 | 307 |
| 2022/5/26 | 12 | 219 | 231 |
| 2022/5/27 | 21 | 131 | 152 |
| 2022/5/28 | 11 | 93 | 104 |
| 2022/5/29 | 4 | 61 | 65 |
| 2022/5/30 | 7 | 22 | 29 |
| 2022/5/31 | 4 | 10 | 14 |
| Total | 35,465 | 591,341 | 626,806 |

Data from <http://www.scio.gov.cn/xwfb/dfxwfb/gssfbh/sh_13834/>.

#### Supplementary Table 2. Containment and health index during the Shanghai Omicron BA.2 wave

| Date | Containment and health index (CHI) |
| --- | --- |
| 2022/3/1 | 64.88 |
| 2022/3/2 | 64.88 |
| 2022/3/3 | 64.88 |
| 2022/3/4 | 64.88 |
| 2022/3/5 | 64.88 |
| 2022/3/6 | 64.88 |
| 2022/3/7 | 64.88 |
| 2022/3/8 | 64.88 |
| 2022/3/9 | 64.88 |
| 2022/3/10 | 68.45 |
| 2022/3/11 | 68.45 |
| 2022/3/12 | 72.02 |
| 2022/3/13 | 74.4 |
| 2022/3/14 | 74.4 |
| 2022/3/15 | 74.4 |
| 2022/3/16 | 76.19 |
| 2022/3/17 | 79.76 |
| 2022/3/18 | 79.76 |
| 2022/3/19 | 79.76 |
| 2022/3/20 | 79.76 |
| 2022/3/21 | 79.76 |
| 2022/3/22 | 79.76 |
| 2022/3/23 | 81.55 |
| 2022/3/24 | 81.55 |
| 2022/3/25 | 81.55 |
| 2022/3/26 | 81.55 |
| 2022/3/27 | 81.55 |
| 2022/3/28 | 95.83 |
| 2022/3/29 | 95.83 |
| 2022/3/30 | 95.83 |
| 2022/3/31 | 95.83 |
| 2022/4/1 | 95.83 |
| 2022/4/2 | 95.83 |
| 2022/4/3 | 95.83 |
| 2022/4/4 | 98.21 |
| 2022/4/5 | 98.21 |
| 2022/4/6 | 98.21 |
| 2022/4/7 | 98.21 |
| 2022/4/8 | 98.21 |
| 2022/4/9 | 98.21 |
| 2022/4/10 | 98.21 |
| 2022/4/11 | 97.02 |
| 2022/4/12 | 97.02 |
| 2022/4/13 | 97.02 |
| 2022/4/14 | 97.02 |
| 2022/4/15 | 97.02 |
| 2022/4/16 | 94.64 |
| 2022/4/17 | 94.64 |
| 2022/4/18 | 94.64 |
| 2022/4/19 | 94.64 |
| 2022/4/20 | 94.64 |
| 2022/4/21 | 94.64 |
| 2022/4/22 | 94.64 |
| 2022/4/23 | 94.64 |
| 2022/4/24 | 94.64 |
| 2022/4/25 | 94.64 |
| 2022/4/26 | 94.64 |
| 2022/4/27 | 94.64 |
| 2022/4/28 | 94.64 |
| 2022/4/29 | 94.64 |
| 2022/4/30 | 94.64 |
| 2022/5/1 | 94.64 |
| 2022/5/2 | 94.64 |
| 2022/5/3 | 94.64 |
| 2022/5/4 | 94.64 |
| 2022/5/5 | 94.64 |
| 2022/5/6 | 94.64 |
| 2022/5/7 | 94.64 |
| 2022/5/8 | 94.64 |
| 2022/5/9 | 94.64 |
| 2022/5/10 | 94.64 |
| 2022/5/11 | 94.64 |
| 2022/5/12 | 94.64 |
| 2022/5/13 | 94.64 |
| 2022/5/14 | 94.64 |
| 2022/5/15 | 94.64 |
| 2022/5/16 | 94.64 |
| 2022/5/17 | 94.64 |
| 2022/5/18 | 94.64 |
| 2022/5/19 | 94.64 |
| 2022/5/20 | 94.64 |
| 2022/5/21 | 94.64 |
| 2022/5/22 | 94.64 |
| 2022/5/23 | 94.64 |
| 2022/5/24 | 94.64 |
| 2022/5/25 | 94.64 |
| 2022/5/26 | 94.64 |
| 2022/5/27 | 94.64 |
| 2022/5/28 | 94.64 |
| 2022/5/29 | 94.64 |
| 2022/5/30 | 94.64 |
| 2022/5/31 | 94.64 |

Data from https://github.com/OxCGRT/covid-policy-tracker.

#### Supplementary Table 3. Parameters in the regression model

| Parameters | Period of effective reproduction number  selected for analysis | | |
| --- | --- | --- | --- |
|  | 2022-3-2 to 5-24  (12 weeks) | 2022-3-2 to 5-17  (11 weeks) | 2022-3-2 to 5-10  (10 weeks) |
| $\boldsymbol{\alpha}$ | -35.71  (-36.78, -34.77) | -36.20  (-37.11, -35.37) | -37.62  (-38.52, -36.81) |
| $\boldsymbol{\varepsilon}$ | -0.0071  (-0.0099, -0.0047) | -0.0069  (-0.0096, -0.0045) | -0.0065  (-0.0092, -0.0040) |
| $\boldsymbol{ln(}\boldsymbol{R}_{\boldsymbol{0}}\boldsymbol{S}_{\boldsymbol{0}}\boldsymbol{)}$ | 1.15  (0.93, 1.40) | 1.14  (0.92, 1.38) | 1.11  (0.88, 1.35) |
| *R-squared* | 0.975  (0.964, 0.985) | 0.974  (0.962, 0.984) | 0.974  (0.961, 0.986) |

#### Supplementary Figure 1. Policy indicators related to policies

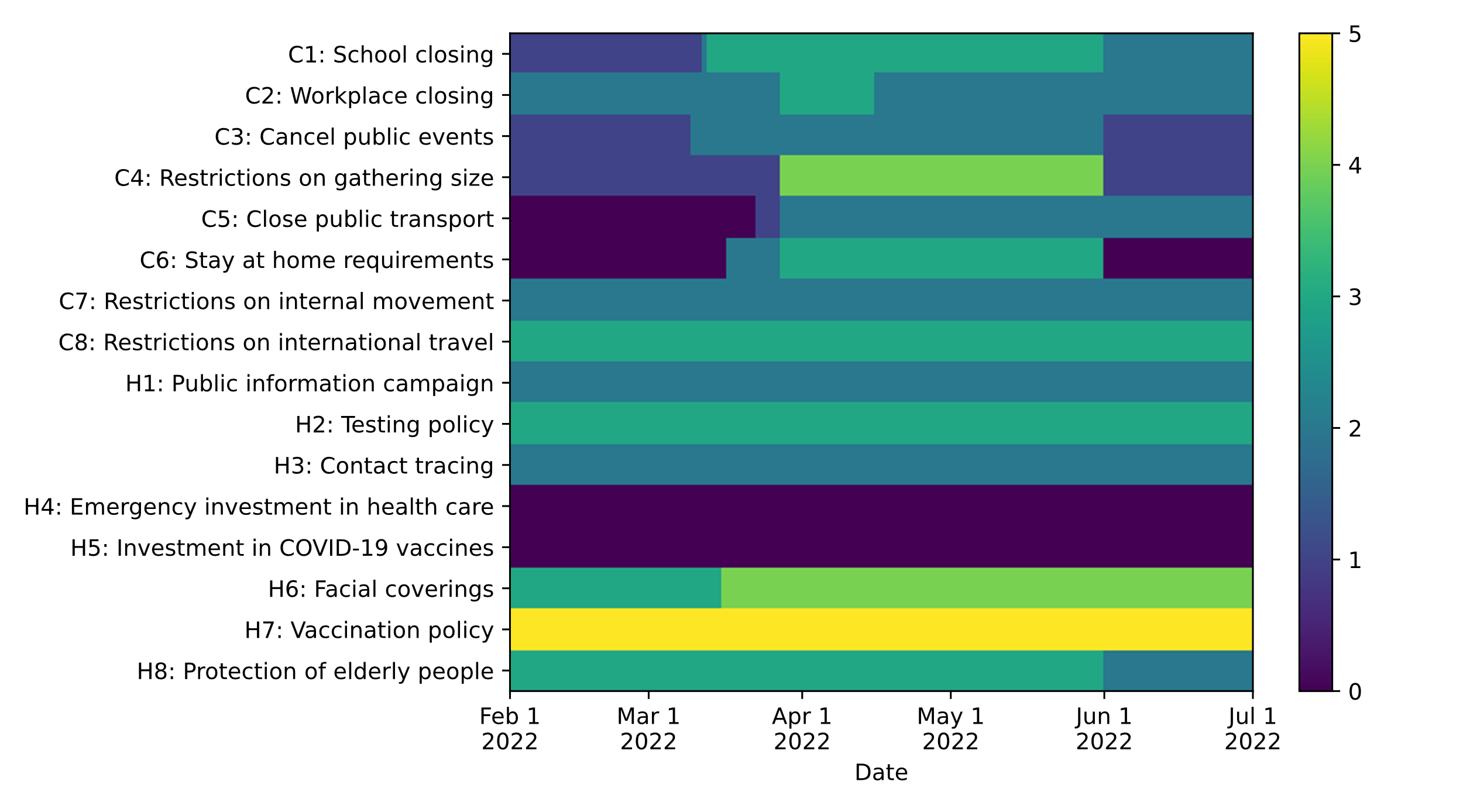

Supplementary Figure 1. Containment and closure policies (C1-C8) and health system policies (H1-H8) over time during the Omicron BA.2 outbreak in Shanghai.

#### Supplementary Figure 2. Epidemic curves by date of reporting and sample collection

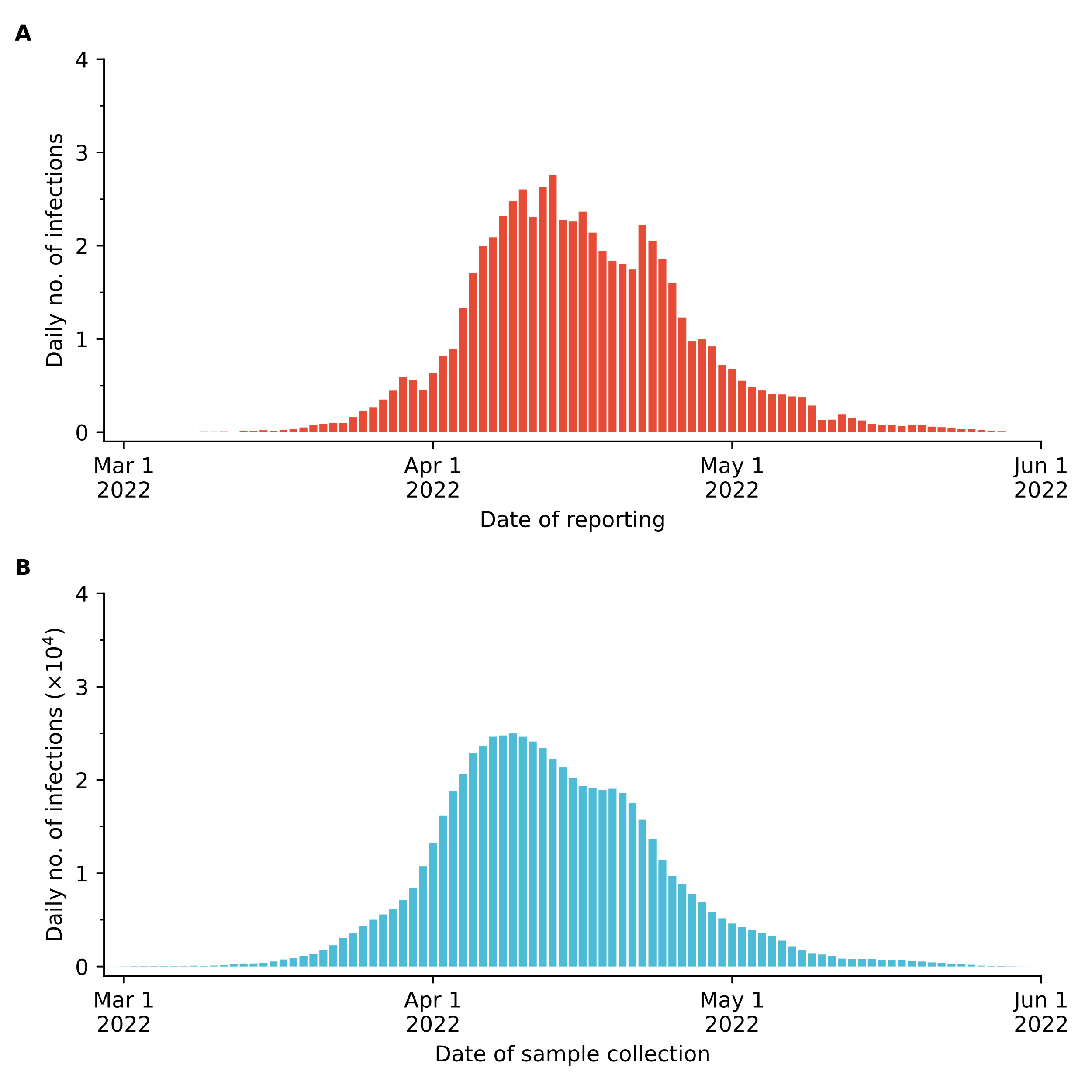

Supplementary Figure 2. **A** Daily number of newly reported infections by date of reporting. **B** Daily number of newly reported infections by date of sample collection. Delays of 2 days, 3 days, and 2 days between the sampling date and the reporting date for the periods before March 15, between March 16 and May 14, and after May 15 are considered to generate the sampling date for each infection.

#### Supplementary Figure 3. Sensitivity analysis of the effective reproduction number

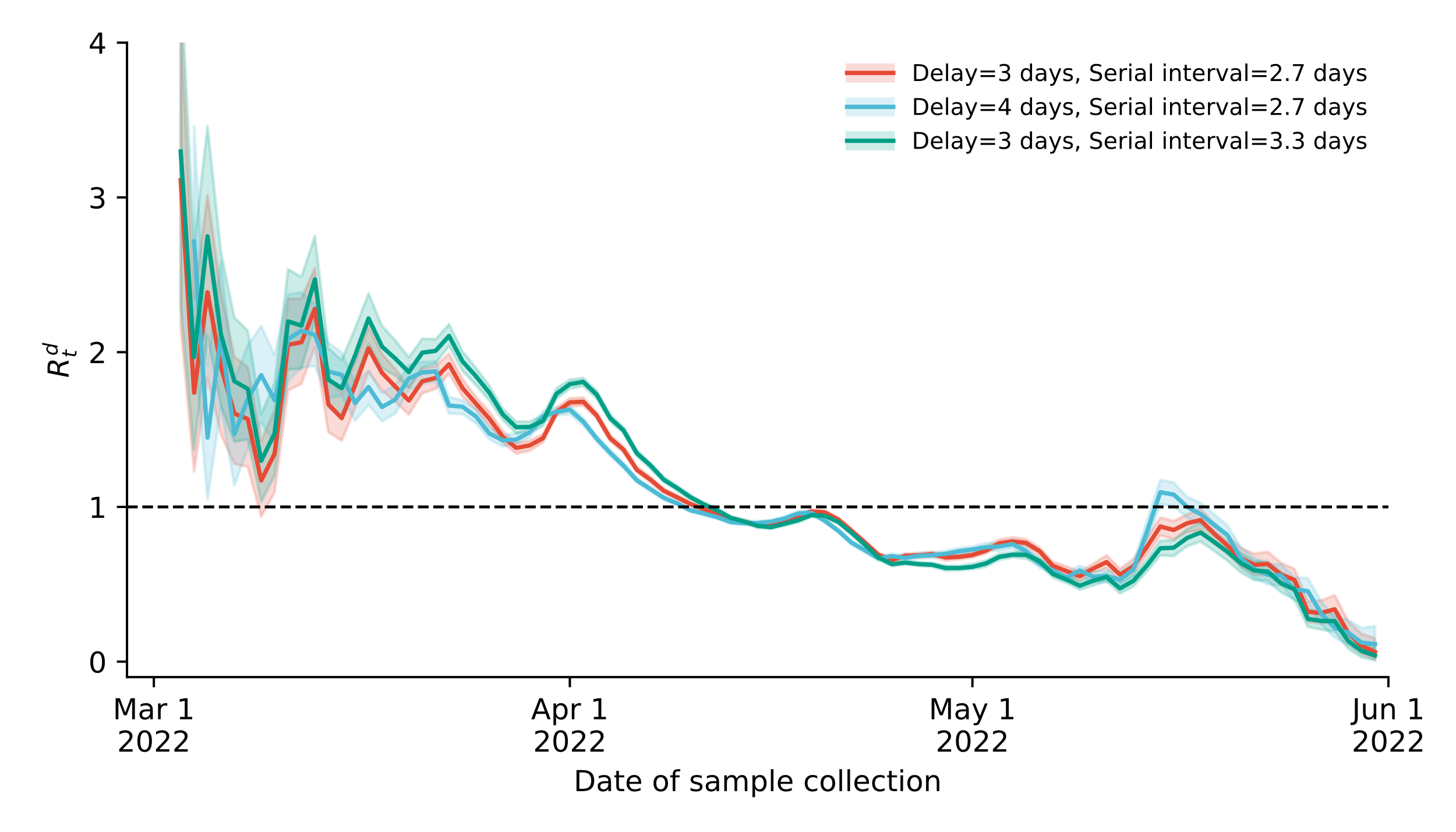

Supplementary Figure 3. Delays of 2 days, 3 days (red curve)/4 days (blue curve), and 2 days between the sampling date and the reporting date for the periods before March 15, between March 16 and May 14, and after May 15 are considered to generate the sampling date for each infection. A gamma-distributed serial interval with a mean of 2.7 days (red curve) and 3.3 days (green curve) are used in the Bayesian method to estimate the effective reproduction number. Line and shaded area: mean and 95% CI. The horizontal line represents the epidemic threshold for the reproduction number.

#### Supplementary Figure 4. Lengthening the duration of dataset to 12 weeks

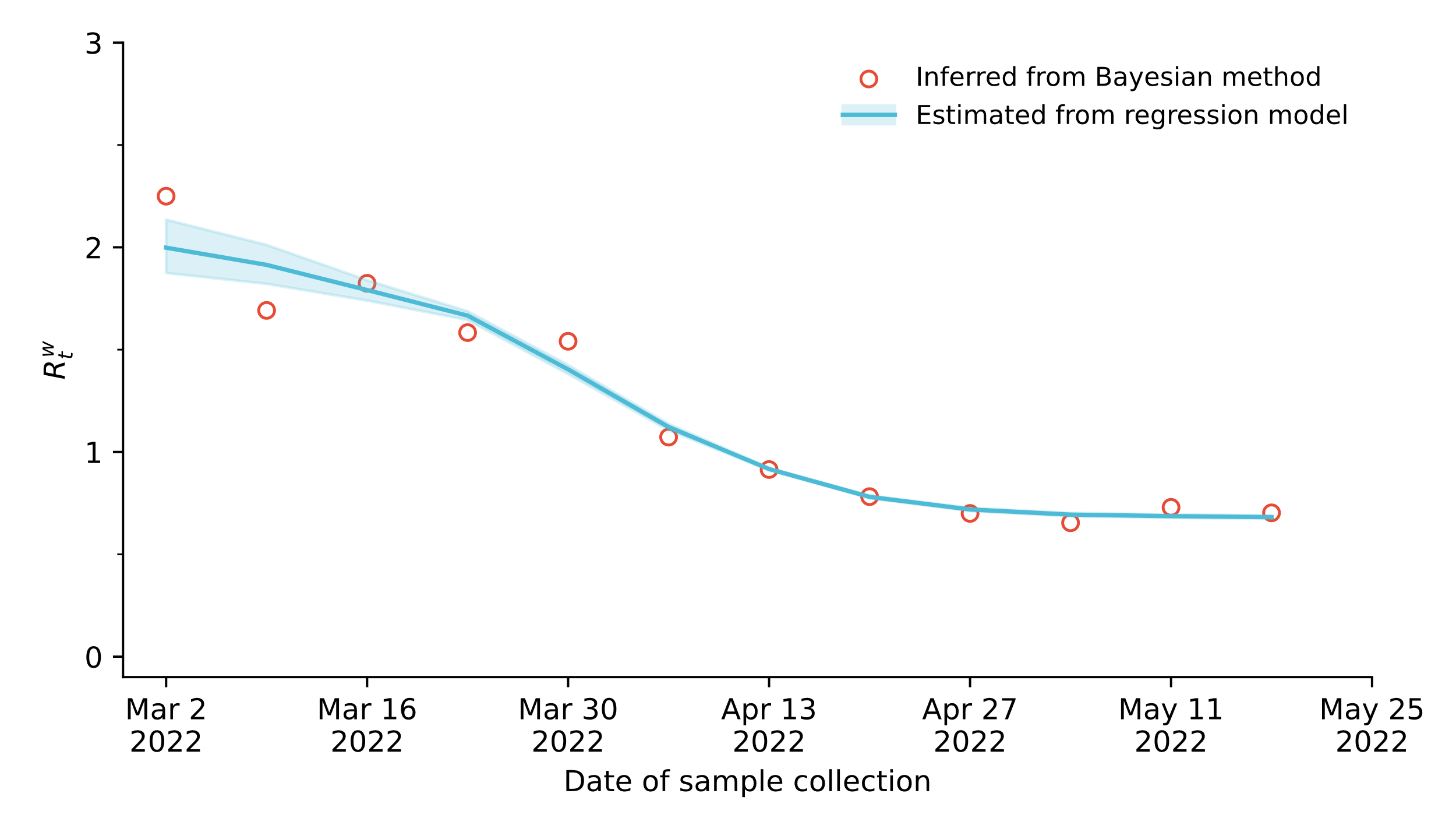

Supplementary Figure 4. Weekly effective reproduction number as estimated from our Bayesian approach (red circles) and as estimated from our regression model (blue line). A 12-week dataset (from March 2 to May 24, 2022) is selected for analysis. The first day is employed to represent the perspective week. Blue line and shaded areas: mean and 95% CI of 100 simulations.

#### Supplementary Figure 5. Shorting the duration of dataset to 10 weeks

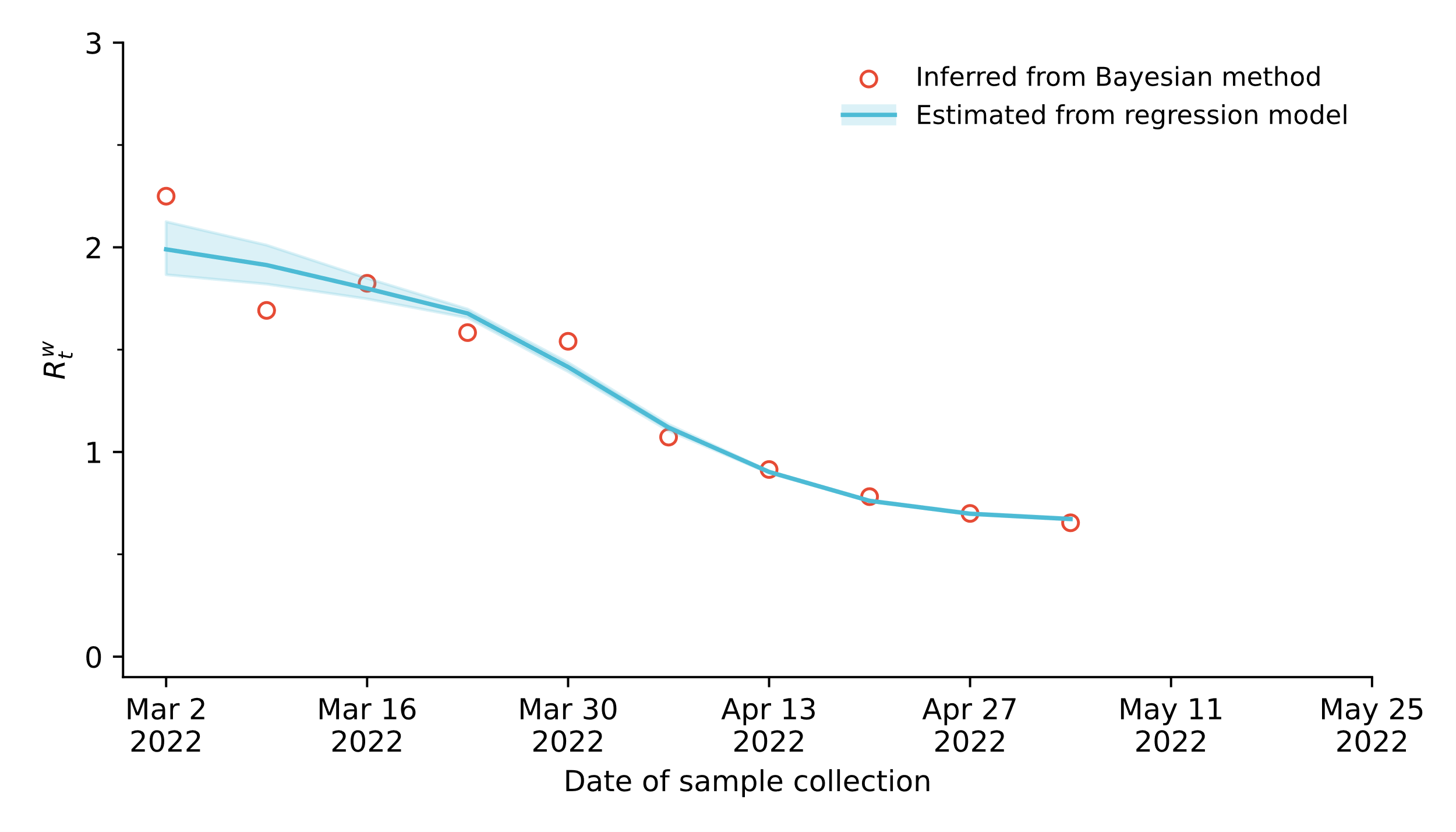

Supplementary Figure 5. Weekly effective reproduction number as estimated from our Bayesian approach (red circles) and as estimated from our regression model (blue line). A 10-week dataset (from March 2 to May 10, 2022) is selected for analysis. The first day is employed to represent the perspective week. Blue line and shaded areas: mean and 95% CI of 100 simulations.
